# Robot-assisted rehabilitation supports cortical network reorganization after nerve transfer surgery to treat chronic, complete cervical spinal cord injury

**DOI:** 10.64898/2026.05.26.26353736

**Authors:** Amanda Bernstein, Justin M. Brown, Kathleen Friel, Edmund Hollis

## Abstract

Recovery of hand and arm function is critical for improving quality of life in individuals with tetraplegia due to spinal cord injury (SCI). Nerve transfer procedures can restore meaningful hand and arm function in chronic SCI, yet postoperative outcomes vary widely. We conducted a prospective, single-arm, open-label trial to assess the impact of intensive, robot-assisted rehabilitation training on functional recovery and cortical reorganization following nerve transfer. The primary endpoint was assessment of hand and arm function measured by the Box and Blocks Test.

We report the results from three participants, AIS A at enrollment, who completed six weeks of intensive robotic training at least 1 year after nerve transfer surgery (NCT04041063). All participants demonstrated minimally important difference improvements in at least one secondary clinical outcome. These improvements were accompanied by cortical reorganization measured by transcranial magnetic stimulation motor mapping, indicating integration of the newly established peripheral motor pathways. No serious adverse events related to surgery or rehabilitation occurred.

Although recruitment was limited by the COVID-19 pandemic and precludes definitive conclusions regarding efficacy, these findings suggest that standardized, intensive robotic rehabilitation may enhance functional outcomes after nerve transfer surgery for chronic tetraplegia.

## Introduction

An estimated 253,000 to 378,000 individuals in the United States live with the long-term effects of spinal cord injury (SCI), over more than half have cervical-level injuries that result in tetraplegia ^1^. Loss of hand and arm function profoundly limits independence, affecting the ability to perform essential activities of daily living such as feeding, dressing, and grooming ^2,3^. For people with tetraplegia, restoring upper-extremity function is consistently identified as the highest rated priority, given its direct impact on quality of life and autonomy ^4^. Yet treatment options remain limited, particularly for those with complete injuries classified AIS A on the American Spinal Injury Association (ASIA) Impairment Scale. AIS A injuries are the most common at discharge (40.9%) and account for 33.9% of individuals with tetraplegia ^5^, rendering many ineligible for recent transcutaneous stimulation approaches aimed at functional restoration. Even modest improvements in hand or arm movement can meaningfully enhance daily function for this population.

Nerve transfer surgery has emerged as an important reconstructive option for individuals with mid-to-low cervical injury ^6–8^, a population that represents nearly one-third of those living with chronic SCI ^1,9^. By coaptating intact, functionally redundant donor nerves to recipient nerves lacking supraspinal control below the level of injury, nerve transfer can restore volitional control without sacrificing critical spared function ^10^. Donor nerves can re-innervate multiple muscles, potentially expanding functional motor networks ^11^. Although an increasing number of successful procedures have been reported ^8,12,13^, recovery of dexterity following nerve transfer remains highly variable across individuals ^10^.

Intensive, target-specific training may help reduce this variability and improve outcomes. In individuals with chronic tetraplegia, six weeks of intensive, robot-assisted upper-extremity training has been shown to improve kinetic aspects of hand and arm movement ^14^. Robotic systems allow high-throughput, high-intensity practice with precise, quantitative assessment of movement kinematics, making them well suited for standardized post-surgical rehabilitation ^15^. These features suggest a possible role for robotic training in enhancing outcomes after nerve transfer, although this has not yet been systematically evaluated.

Recovery of dexterity after SCI also depends critically on plasticity within sensorimotor cortical networks. Shortly after injury, cortical representations of body regions above the lesion expand into areas corresponding to the impaired body regions ^16,17^. As motor function recovers, the cortical territory responsive to the recovered movement increases ^18^. Similar training-dependent cortical remodeling has been documented in animal models ^19,20^, and lesioning of the reorganized motor cortex reverses behavioral gains, underscoring its causal role in functional recovery ^21^. However, the capacity of the human motor cortex to reorganize and incorporate the novel peripheral circuits created by nerve transfer, and the potential for rehabilitation to shape this reorganization, remain unknown.

We therefore tested the hypothesis that intensive robot-assisted training would support both 1) improved recovery of hand and arm function and 2) cortical reorganization reflecting incorporation of the newly established peripheral circuits after nerve transfer surgery. Here, we report the findings from three adults with chronic tetraplegia who completed six weeks (18 sessions) of intensive robotic rehabilitation at least 12 months after nerve transfer (NCT04041063). Rehabilitation was delivered using the InMotion ARM/HAND robot (Bionik, Watertown, MA), a device previously validated for reducing upper-extremity impairment in people with SCI ^14^, stroke ^22–25^, and cerebral palsy ^26^.

### Study design

This case is part of clinical trial NCT04041063, approved by the BRANY Institutional Review Board, and listed on Clinicaltrials.org before participant enrollment. The participants provided written consent for all study procedures. Clinical outcome measures, including assessments of hand and arm function, kinematic outcomes, and transcranial magnetic stimulation (TMS) mapping of motor cortex, were performed once prior to surgery, twice at a minimum of one year after surgery (∼1 week apart), and once again after six weeks of robotic training. Details on outcome measures are provided below.

Transcutaneous stimulation with a SunStim (SunMed, Grand Rapids, MI) peripheral nerve stimulator at the 100 Hz tetanus with maximum strength was used pre-operatively to confirm functional innervation of target muscles during initial screening procedure. Surgeries were performed by Dr. Justin Brown at the Massachusetts General Hospital Paralysis Center.

## Methods

### Participant recruitment

The trial included adult participants aged 18-65 years who had sustained a traumatic cervical (C4-C7) SCI more than 6 months before their enrollment. Only participants who responded to pre-operative nerve or direct muscle stimulation testing of the target muscles were candidates for nerve transfer surgery and therefore could be enrolled. All participants were capable of providing written informed consent. Participants were excluded from the study if they had any potential risk factors for brain stimulation, such as a metal implant in their head, or if they had a cognitive deficit that would prevent them from following the training instructions on the robot. In total, 9 participants were assessed for eligibility (Figure 1). Of those, 5 were excluded due to not meeting the inclusion criteria (*n* = 3) or declining to participate further in the study (*n* = 2). Those that declined to participate did so out of travel concerns during the COVID-19 pandemic. Four people received the nerve transfer surgery, and three completed 6 weeks of robotic training. One person was unable to complete all 18 study visits and dropped out of the study. Out of the three people that completed the 6 weeks of robotic training, one person elected to return for a one-year follow-up evaluation.

**Figure 1.**
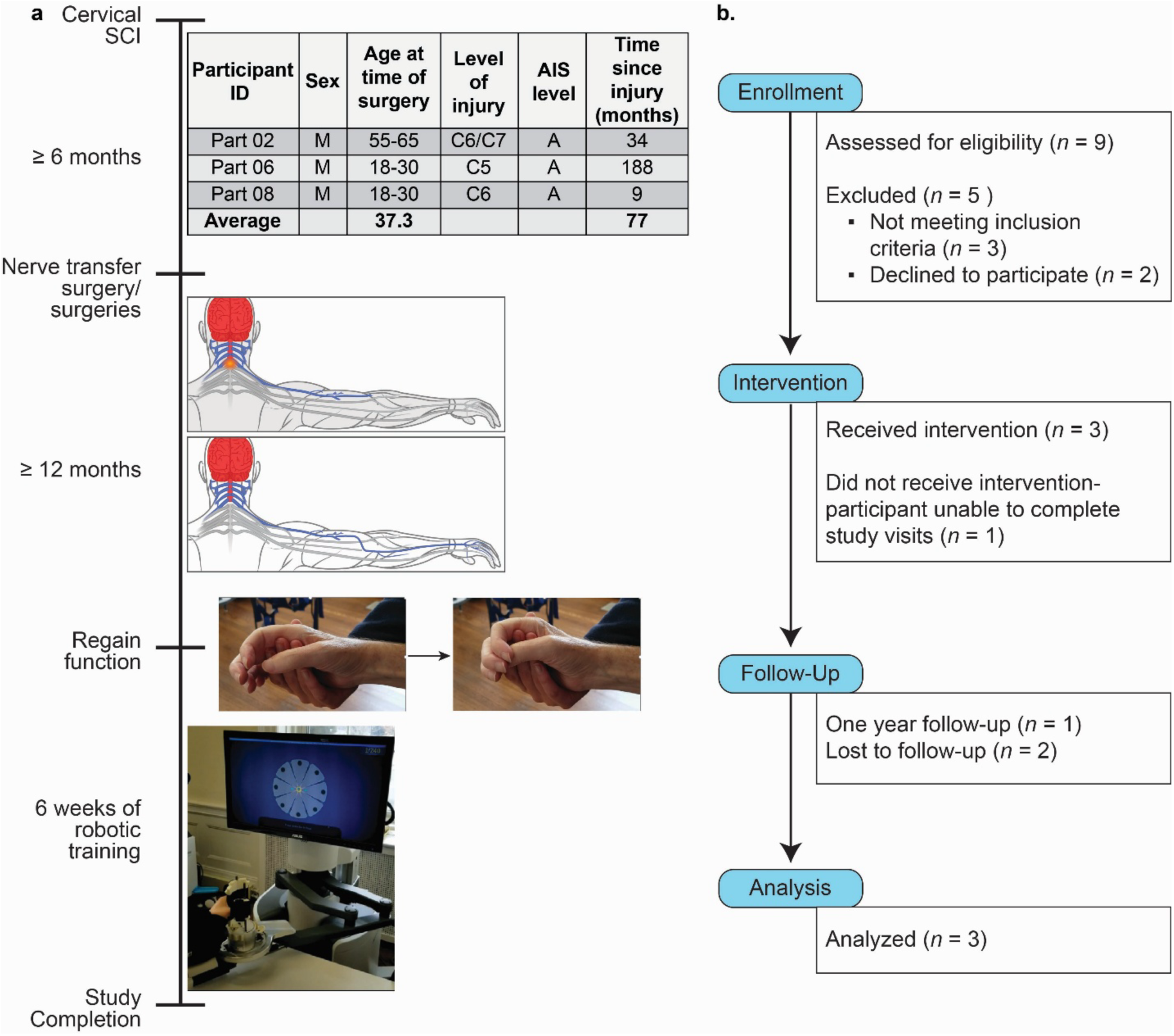
Study timeline and participant flow for trial NCT04041063. This pilot trial examined the effects of robotic-assisted rehabilitation on dexterous hand movements and cortical motor map changes in tetraplegic individuals following nerve transfer surgery. Participants were screened and enrolled after a baseline assessment. Individuals were excluded if they did not meet eligibility criteria or declined participation, including for COVID-19 related travel concerns. Enrolled participants received nerve transfer surgery and, at least one year later, completed 6 weeks of robotic training. Participants were considered to have completed the study after finishing all rehabilitation sessions and post-training assessments.

### Participant details

#### Participant 2

a 55-65-year-old male presented with C6-C7 American Spinal Injury Association Impairment Scale (AIS) A at 34 months after traumatic SCI. His recovery had plateaued, and he had previously completed six weeks of robotic rehabilitation on the InMotion ARM/HAND robot as part of study NCT03555838. He exhibited volitional control of the biceps, elbow extensors and flexors, and wrist extensors, but limited hand control was observed. Participant 2 initially enrolled for right arm treatment, adding left arm treatment at a later date, resulting in an absence of baseline mapping and robotic assessments for his left arm. Therefore, his left arm data is provided in a supplement (Tables 2, 4, 6, 8, 10 and Figures S3-5) and not included in our main analysis. Nerve transfer surgery was performed on the right arm three years after his injury and on the left arm at eight months after the right arm surgery. Right arm surgery: supinator was transferred to the anterior interosseus nerve (AIN). Left arm surgery: pronator teres nerve was transferred to AIN. In addition, supinator and a branch from extensor carpi radialis brevis (ECRB) were transferred to posterior interosseus nerve (PIN).

**Table 1.**
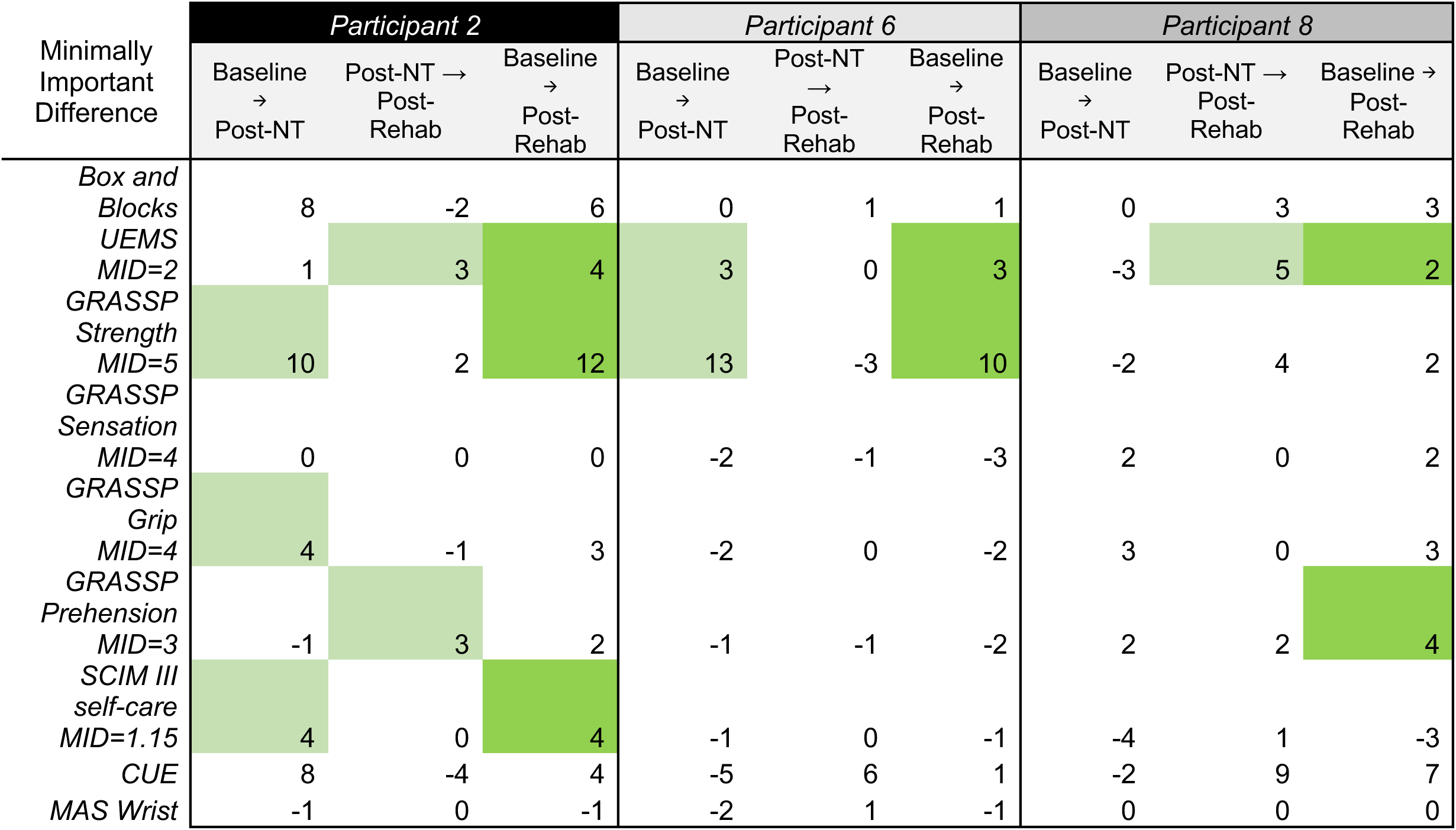
Summary of responder status across clinical outcomes. All participants met responder criteria for at least one strength-related test. Participants were classified as responders for a given outcome if their change in score met or exceeded the minimally important difference (MID), calculated as the change in score between the Baseline evaluation and the end of the robotic training period. Strength-domain outcomes included the GRASSP Strength score (MID = 5-point increase) and the UEMS (MID = 2-point increase). Color shading indicates responder status for each outcome.

**Table 2.** Persistence of improved clinical and kinematic measures of hand function at follow-up. Participant 2 showed stability and mild improvements at 19 months (right arm) and 9 months (left arm) after completing 6 weeks of robotic training. ^†^Smaller values indicate better performance. *_ý_* Indicates measures that had improved after 6 weeks of robotic training.

| Outcome<br>Measures | Participant 2 Left Hand |  | Participant 2 Right Hand |  |
| --- | --- | --- | --- | --- |
|  | Post-Rehab | 9 months<br>Post-Rehab | Post-Rehab | 19 months<br>Post-Rehab |
| *Box and Blocks* | 4 | 6 | 23 | 20 |
| *UEMS<sub>ψ</sub>* | 13 | 15 | 18 | 16 |
| *GRASSP Strength<sub>ψ</sub>* | 18 | 20 | 25 | 22 |
| *GRASSP Sensation* | 4 | 8 | 7 | 7 |
| *GRASSP Grip* | 3 | 3 | 5 | 8 |
| *GRASSP Prehension<sub>ψ</sub>* | 3 | 4 | 10 | 14 |
| *SCIIM self-care* | 8 | 5 | 9 | 5 |
| *CUE* | 34 | 32 | 38 | 42 |
| *Hand Release AROM* | 4.17 cm | 4.31 cm | 4.08 cm | 4.17 cm |
| *Hand Grasp AROM<sup>†</sup>* | 4.4 cm | 4.44 cm | 4.23 cm | 4.28 cm |
| *Hand Release Stabilization<sub>ψ</sub>* | 4.42 cm | 5.15 cm | 4.5 cm | 4.37 cm |
| *Hand Release Force<sub>ψ</sub>* | 1 N | 5.1 N | 2 N | 0.2 N |
| *Hand Grasp Stabilization<sup>†</sup>* | 7.03 cm | 6.75 cm | 6.68 cm | 6.41 cm |
| *Hand Grasp Force* | 9 N | 11.8 N | 12.4 N | 14.9 N |
| *Circle Size<sub>ψ</sub>* | 88.85 cm | 88.79 cm | 88.55 cm | 89.13 cm |
| *Circle Symmetry* | 0.858 | 0.879 | 0.906 | 0.932 |
| *Target Accuracy* | 0.85 cm | 0.87 cm | 0.91 cm | 0.9 cm |
| *Movement Efficiency* | 91% | 91% | 88% | 89% |
| *Isometric Hold Distance<sup>†</sup><sub>ψ</sub>* | 3.3 cm | 2.71 cm | 4.11 cm | 3.03 cm |
| *Isometric Hold Force<sub>ψ</sub>* | 24.6 N | 25.5 | 21.3 N | 24.1 N |
| *Displacement Against Resistance* | 13.07 cm | 13.13 cm | 13.09 cm | 13.08 cm |
| *Resistance Force* | 26.6 N | 26.7 N | 26.8 N | 26.6 N |

#### Participant 6

a 18-30-year-old male presented with a C4-C5 fracture (AIS A) due to traumatic SCI 15 years earlier. He reported bilateral weakness in hands and arms, and though he performs exercises at home, has not regained any new function. The first nerve graft was a bilateral brachialis to sural graft completed approximately 15 years after his injury on the left arm. Then, a sural graft to AIN hook up with the supinator connected to the PIN was completed 7 months later.

#### Participant 8

a 18-30-year-old male presented with C6 AIS A that converted to C4 AIS A at 9 months after traumatic SCI. He reported bilateral weakness in hands and arms, with some improvements in strength and movement in the left arm. He exhibited volitional control of biceps, but limited control of the wrist extensors and flexors, elbow extensors, and hand movements. The first nerve transfer surgery was performed on the left arm nine months after his injury, then a second nerve transfer was performed 6 months after the first. A tendon transfer was performed 15 months after the last nerve transfer surgery. For the first surgery on the left arm surgery, the teres minor was transferred to the long head triceps. In addition, the brachialis was transferred to the sural nerve graft, which was then connected to the AIN and flexor digitorum superficialis (FDS) during the second nerve transfer surgery. For the tendon transfer surgery, the flexor carpi radialis (FCR) was transferred to the extensor pollicis longus (EPL) and the brachioradialis was transferred to the flexor pollicis longus (FPL).

### Intensive robotic rehabilitation

At least one year post-surgery, participants, seated in their own wheelchairs, completed 18 sessions (3x/week, 6 weeks, 90 min) of interactive hand robotic training on the InMotion ARM/HAND robot. In each session, participants performed a total of 1,070 movement repetitions using one arm, resulting in over 18,000 repetitions over the six-week period focused on the biceps, triceps, and flexors/extensors of the hand that produce a cylindrical grip. The interactive robotic features involve a visuomotor task, moving the robotic manipulandum according to targets on a computer screen mounted at eye level. The surgically treated arm was abducted, and the forearm supported, with the hand grasping contoured finger and thumb grips. Velcro straps lightly held the forearm and fingers secure, preventing the potential use of tenodesis to open and close the hand. The robot provides adaptive assistance in gross grasp and release motions and support for functional reach-grasp-release movements (Masia et al., 2007). The training consisted of 80 repetitions of grasping and release in place, 480 repetitions of moving to a target then grasping and releasing, and 480 repetitions of squeezing, dragging, and releasing an object on the screen at each target. Before and in-between each training module, the participants had to perform 5 unassisted grasps and release in place.

### Outcome measures

Clinical outcome measures were taken prior to surgery, at least 12 months after nerve transfer surgery, and again after six weeks of robotic training was completed. For Participant 2, clinical outcome measures were taken 12 months after nerve transfer surgery on the right hand, and 14 months after nerve transfer surgery on the left hand. A follow-up evaluation was performed 19 months (9 months left hand) after completed robotic training of the right hand to determine the persistence of robotic training effects. For Participant 6, clinical outcome measures were taken 16 months after the second nerve transfer. For Participant 8, clinical outcome measures were taken 21 months after nerve transfer, 6 months after tendon transfer. Hand and arm function was tested bilaterally using the Box and Blocks test of manual dexterity. Participants were required to grasp and transport as many 2.5cm^3^ blocks as possible from one box to another, over a barrier, in one minute. Clinical motor strength was tested by the International Standards for Neurological Classification of Spinal Cord Injury Upper Extremity Motor Score (UEMS). UEMS grades motor strength on the American Spinal Injury Association (ASIA) scale from 0 to 5 for each key muscle group (elbow flexors, elbow extensors, wrist extensors, finger flexors, finger extensors). This sum score ranges from 0 (paralyzed) to 25 (normal) in each limb.

Other functional assessments included the Graded Redefined Assessment of Strength Sensibility and Prehension V2 (GRASSP) ^27^ assessment of specific impairment of the upper limb in tetraplegia. GRASSP V2 is an interval scale with four parts that score strength (scored 0-50 for each arm), sensation (scored 0-12 for each hand), prehension ability (scored 0-12 for each hand), and prehension performance (scored 0-20 for each hand). Spasticity was measured by the Modified Ashworth Scale (MAS) ^28^, a six point ordinal scale of elbow spasticity.

To assess quality of life, the participants completed questionnaire-based assessments: the Spinal Cord Independence Measure (SCIM III) ^29^ to measure their ability to perform everyday tasks and the Capabilities of Upper Extremity Instrument (CUE) to assess upper extremity functional limitations. SCIM III was used to provide quantitative assessments of 19 tasks in 16 categories (score range 0-100); all activities of daily living, grouped into four areas of function (subscales): Self-Care (scored 0–20), Respiration and Sphincter Management (0–40), Mobility in Room and Toilet (0–10), Mobility Indoors and Outdoors (0–30). In this report only the Self-Care sub-score of the SCIM III questionnaire was reported. CUE has 4 main domains; perceived ability to reach or lift, perceived ability to pull and push with arms, perceived ability to move and position the arm/wrist, and perceived ability to use hands/fingers. Each question is scored on a scale of 0 (unable to complete) to 4 (no difficulty) for both arms separately as well as a couple of questions about bimanual tasks for a total score of 128 (score range 0-60 for each arm). In this report only the scores for individual arms (score range 0-60) were reported.

In addition to providing training on visuomotor tasks, the InMotion ARM/HAND robot was used to obtain sensitive, precise, and reliable quantitative measures of upper extremity movement. We measured the active range of motion of the arm/hand and relative strength of the arm/hand movements bilaterally. We also measured accuracy, smoothness, and efficiency in reaching movements as the participants traced paths in the shapes of stars and circles, as well as the distance that they were able to move against force or resist movement when force was applied. These data identify the components of motor control that underlie arm function, primarily in grasp and release.

### Transcranial Magnetic Stimulation and Motor Mapping

Single-pulse transcranial magnetic stimulation (TMS) was used to map motor cortex contralateral to the arm that received the surgery. The participants were seated in their wheelchair for all mapping sessions. Electromyography (EMG) responses were recorded simultaneously in seven muscles on the tested side of the body: *extensor digitorum communis* (*EDC*), *flexor pollicis longus* (*FPL*), *first dorsal interosseous* (*FDI*), *flexor digitorum profundus* (*FDP*), *extensor carpi radialis* (*ECR*), *biceps brachii* (*BIC*), and *soleus*. Bipolar electrodes were used, with one electrode placed on the muscle belly, and the other placed on the tendon. EMG responses were recorded using the NeuroPrax EXG system (NeuroConn, Germany). Each time a TMS pulse was delivered, a trigger was sent to the NeuroPrax to mark in the EMG files.

A Magstim 200 TMS device (Magstim, UK) with a 70mm figure 8 coil was used to deliver single TMS pulses to the scalp of the participant. A Brainsight neuronavigation system (Rogue Research, Canada) was used to co-register the position of the TMS coil to the participant’s head, using a model MRI, infrared camera, and trackers.

At the beginning of each session, TMS pulses were delivered to the scalp over motor cortex to find the spot that evoked the largest motor evoked potential (MEP) in *EDC*. After this location, the “hotspot,” was found, we next determined the resting motor threshold (RMT). With the participant’s arm relaxed, we stepped down the maximum stimulator output (MSO) by 2% until we identified the minimum MSO needed to produce a 50 μV MEP in at least 6 of 10 trials in *EDC*.

For motor cortex mapping, an 8 cm^2^ grid with 1 cm spacing was superimposed onto the model MRI in Brainsight. The grid was centered on the hotspot. Three stimuli were delivered to each gridpoint at a frequency < 0.2 Hz and an MSO of 1.1x RMT at first session. The grid covered a wide enough area to completely encompass the motor map, as no MEPs were found along the map border.

The RMT, mapping MSO, and grid placement were determined at the first baseline session. In each subsequent mapping session, we measured RMT, but we used the same mapping MSO and grid that were determined at the first session. The Brainsight system enabled us to colocalize the TMS coil position to the participant’s head at each session, which optimized consistency in coil position across sessions.

### Statistical Analysis

A Friedman test with multiple comparisons with an Alpha value of 0.05 for all clinical and kinematic evaluations. The Kruskal-Wallis test was used to assess differences in number of responsive sites and MEP amplitude for each muscle before, at least 12 months after surgery, and after 6 weeks of robotic training. We used nonparametric statistics because the data were not normally distributed.

## Results

### Primary Outcomes

At least one year after nerve transfer surgery, participants showed no significant change in Box and Blocks performance prior to rehabilitation. Following rehabilitation, all participants showed some improvement on the task with one arm (65±18% improvement over pre-surgical baseline) (Figure 2); however, Participant 2 experienced a 73% decline in Box and Blocks performance with their non-dominant left arm (Figure S3), coincident with reduced spasticity. There was significant variability in individuals’ performance on Box and Blocks at all timepoints, limiting interpretation of the findings.

**Figure 2.**
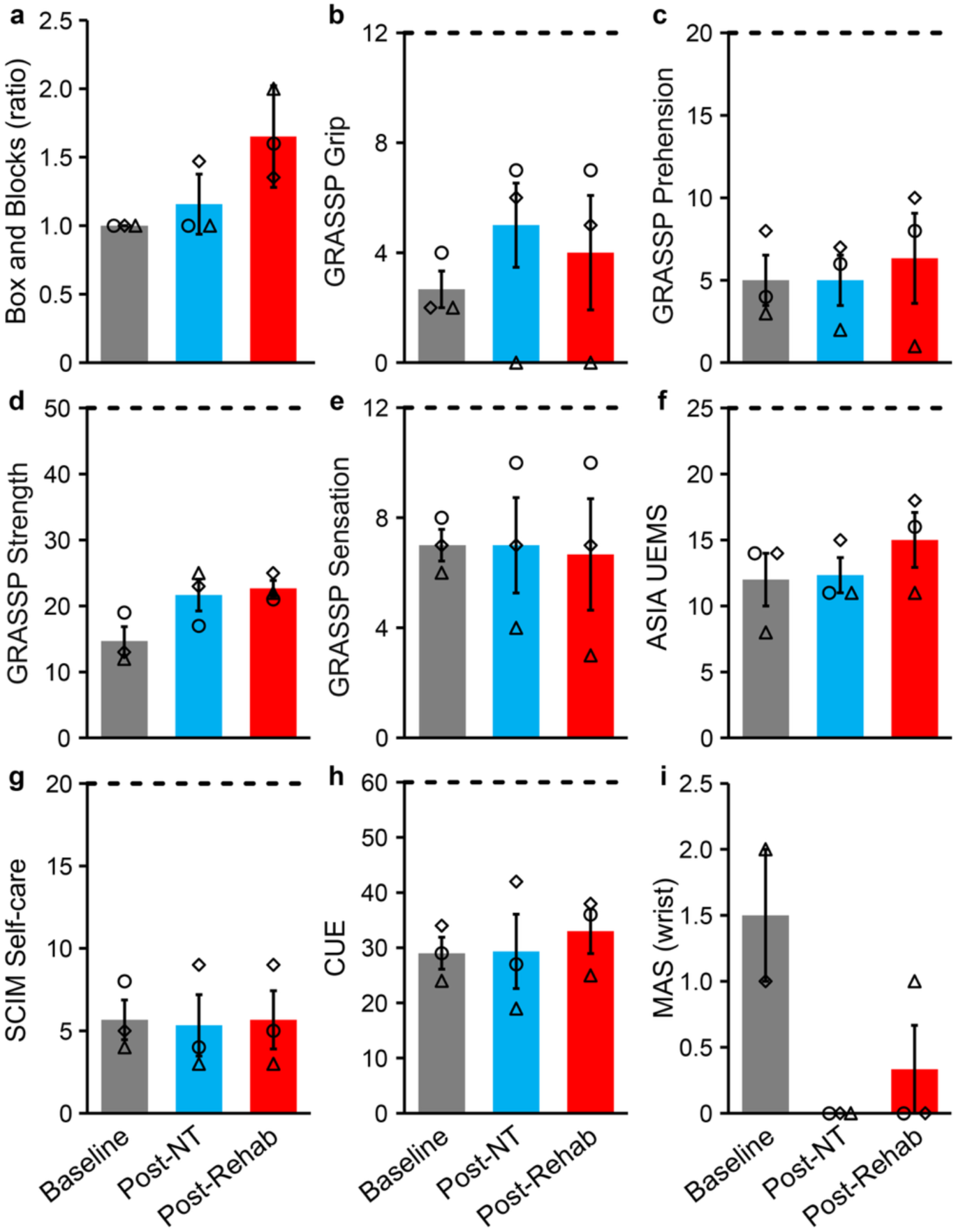
Averaged clinical outcome scores demonstrate mild improvements in hand function following nerve transfer surgery and robotic rehabilitation. (◊ = Participant 2 Right Arm, Δ= Participant 6, ○ = Participant 8). (**a**) Combined treatment of nerve transfer and robotic training contributed to improved Box and Blocks performance, shown as a ratio relative to baseline. (**b-e**) GRASSP measures illustrate the effects of nerve transfer on motor function, with robotic rehabilitation further enhancing strength. (**f**) Combined treatment led to improvements in individual UEMS values. (**g,h**) Quality of life assessments (SCIM III self-care and CUE). (**i**) Wrist spasticity decreased following nerve transfer surgery. Data are presented as mean ± SEM.

### Clinical Outcomes

We analyzed minimally important differences (MIDs) for all clinical outcome measures to identify which changes represented meaningful improvement. Participants were classified as responders if they met the MID for any clinical outcome between enrollment and study completion. All participants demonstrated improvement in at least one strength-related measure: the GRASSP Strength score (MID = 5-point increase) or the UEMS (MID = 2-point increase) (Table 1). Although all participants met responder criteria, average changes across clinical measures of function showed only moderate, non-significant improvement or remained stable relative to pre-surgical baseline (Table S1). Individual participant responses are detailed below.

One participant, Participant 2, showed improvements in their dominant, right hand, but declines in left hand performance (Table S1, S2). Pre-rehabilitation clinical measures were stable as there was limited variability when tested 1 week apart, prior to the start of robotic rehabilitation. Coincident with improved clinical measures of function, Participant 2 noted improvements in quality-of-life SCIM III self-care sub-score and CUE questionnaires after nerve transfer surgery in both arms, while the other two participants saw a slight decrease in the quality-of-life survey scores. Spasticity remained unchanged with a score of 0 or decreased across the study for those exhibiting spasticity at recruitment, Participants 2 (right wrist) and 6 (left wrist). With the loss of spasticity on the left side, Participant 2 also experienced declines in GRASSP prehension performance with the left arm at 14 months post-surgery.

Six weeks of intensive robotic training drove moderate non-significant increases in clinical outcome measures, primarily in prehension capabilities (Figure 2a-f, Table S1). For Participant 2, increases were noted in ASIA UEMS and GRASSP strength and prehension performance values for the right arm and GRASSP grip and prehension performance, as well as SCIM and CUE scores on the left arm. For Participant 6, slight increases were observed in CUE score, while other metrics remained stable (Table S1). For Participant 8, mild increases were observed in ASIA UEMS, GRASSP strength and prehension performance, and CUE tests (Table S1). Both the UEMS and the first GRASSP sub-score measure muscle strength, while the fourth GRASSP sub-score measures prehension performance, which also requires grip strength. Changes in these measures indicate that improvements from robotic training were largely driven by increases in hand strength rather than dexterity or sensation. Increases in quality-of-life survey responses were largely driven by the nerve transfer surgery alone. In summary, despite differences in functional ability prior to nerve transfer surgery, six weeks of intensive robotic training resulted in moderate non-significant improvements in hand strength.

To determine the persistence of robot rehabilitation-mediated effects, Participant 2 was assessed again at nineteen months after completing robotic training on the right arm and nine months on the left. On the right side, he exhibited persistent improvements in GRASSP grip and prehension performance, as well as in CUE score (Table 2). On the left arm, persistent improvements were observed on the Box and Blocks test, ASIA UEMS, and GRASSP strength, sensation, and prehension components (Table 2). Persistent improvements in these measures indicate that the robotic training had mild, but long-lasting effects on hand strength for this participant.

### Kinematics of Arm and Hand Movement

Kinematics of hand movement collected on the InMotion ARM/HAND robot demonstrated effects of both nerve transfer surgery and rehabilitation (Table S3, Figure 3j-l). Overall, no loss of function in the arm was detected due to the nerve transfer surgery (Figure 3, Table S5). Significant improvements in arm function and strength after both the nerve transfer surgery and robotic training were detected as shown by the better performance in the Isometric Hold task from baseline to the study completion (Figure 3d). Nerve transfer surgery alone showed significant improvement in arm flexion against lateral movements (Figure 3m,n). Accuracy and movement efficiency of the arm kinematics also showed non-significant improvement due to the nerve transfer surgery and further improved with robotic training (Figure 3f,i). Participant 2 showed a surgery-mediated increase in grasping active range of motion and grasp force with his right hand, similar to the increase observed in the GRASSP strength sub-score (Table S3); left hand and arm kinematics were not collected prior to surgery. On the right side, six weeks of robotic rehabilitation drove improved strength of hand extensors, with increased hand release force. The left hand showed improved strength after six weeks of intensive robotic training in both the extensors and flexors (Table S6). For both Participant 6 and Participant 8, measures of arm function improved after the surgery, then remained consistent across training sessions. Participant 6 showed a mild improvement in arm movement accuracy and efficiency as well as arm strength after six weeks of intensive robotic training (Table S5). Nerve transfer surgery drove improvements in hand grasp range of motion and force in Participant 6 as well as hand release force (Table S3). Intensive robotic training mildly improved range of motion in both grasping and releasing in addition to grasp force. A decrease in the hand release force and hand grasp force was seen in Participant 8 after the surgery, which was maintained across training.

**Figure 3.**
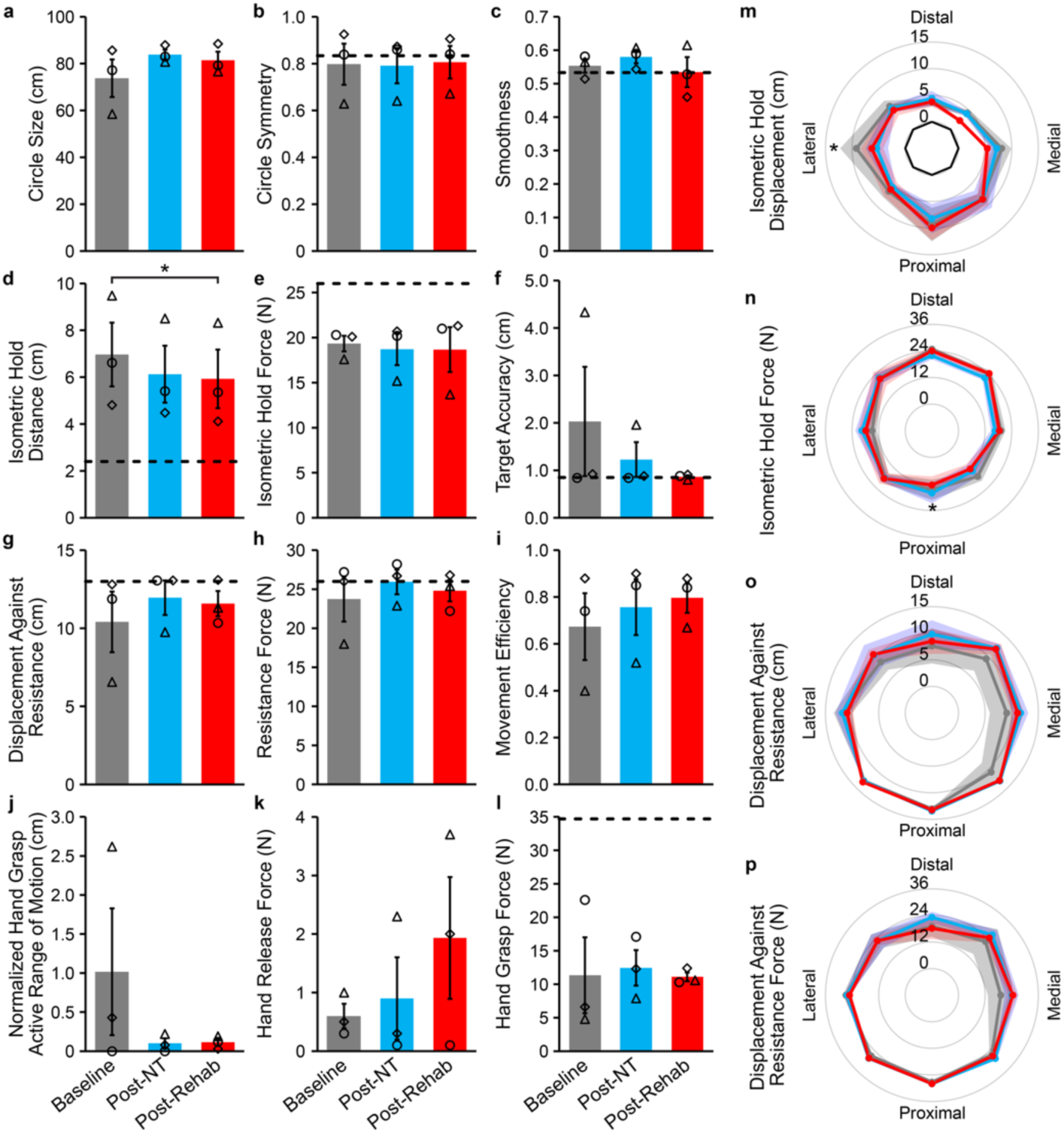
InMotion HAND/ARM robotic measures show preserved function after nerve transfer surgery and improved strength and accuracy of hand and arm movements. (**a-l**) Kinematic measures largely showed minimal changes after nerve transfer surgery and subsequent robotic training; target values represented by dashed lines. (◊ = Participant 2 Right Arm, Δ= Participant 6, ○ = Participant 8). (**d**) Robotic training significantly improved the ability to maintain a static hand position against an external force (Isometric Hold Distance). (**f**) Robotic training increased movement accuracy to the target. (**k**) Robotic rehabilitation supported the restoration of hand release force. (**m-p**) Radar plots depict performance on Isometric Hold Distance/Force and Displacement against Resistance/Force tasks. Significant differences were observed in Isometric Hold Distance from baseline to post-nerve transfer in the lateral direction, and in Isometric Hold Force from post-nerve transfer to post-rehabilitation in the proximal direction. Data are presented as mean ± SEM.

Participant 2 demonstrated persistence of surgery and rehabilitation-mediated effects in both arms during follow-up testing (19 months on right, 9 months on left). In fact, hand grasp force showed continued improvement at follow-up (Table 2). On the right side, finger extensor strength returned to pre-intervention levels, while on the left continued improvement was observed in both finger extensor and flexor strength (Table 2).

### Evoked Motor Maps

Single-pulse TMS revealed non statistically significant cortical motor map remodeling following nerve transfer surgery and subsequent robotic rehabilitation. We assessed changes in both the number of responsive cortical sites and cortical map excitability (MEP size) for six upper extremity muscles contralateral to the arm that underwent nerve transfer. Across participants, robotic rehabilitation drove dynamic changes in muscle-specific map size and excitability, indicating that robotic training contributed to reorganization within motor cortex.

In Participant 2, left motor cortex map sizes remained stable from baseline to 12 months post-surgery for *extensor digitorum communis* (*EDC*), *flexor pollicis longus* (*FPL*), and *extensor carpi radialis* (*ECR*). In contrast, map size decreased for *first dorsal interosseous* (*FDI*), *flexor digitorum superficialis* (*FDS*), and *biceps brachii* (*BIC*). The reduction in *BIC* map size is likely attributable to the nerve transfer, as the donor nerves originally supplied *BIC*. MEP amplitudes did not differ significantly from baseline after surgery. A second post-surgery motor map collected 1 week later confirmed stability: neither the number of responsive sites nor MEP amplitudes differed significantly across the two post-surgery assessments.

After 18 sessions of robotic training, the right arm motor map expanded, with a non-statistically significant increase in the number of responsive sites for all muscles (Table S7, Figure 4). No significant changes in MEP amplitudes emerged in any muscle (Table S9).

**Figure 4.**
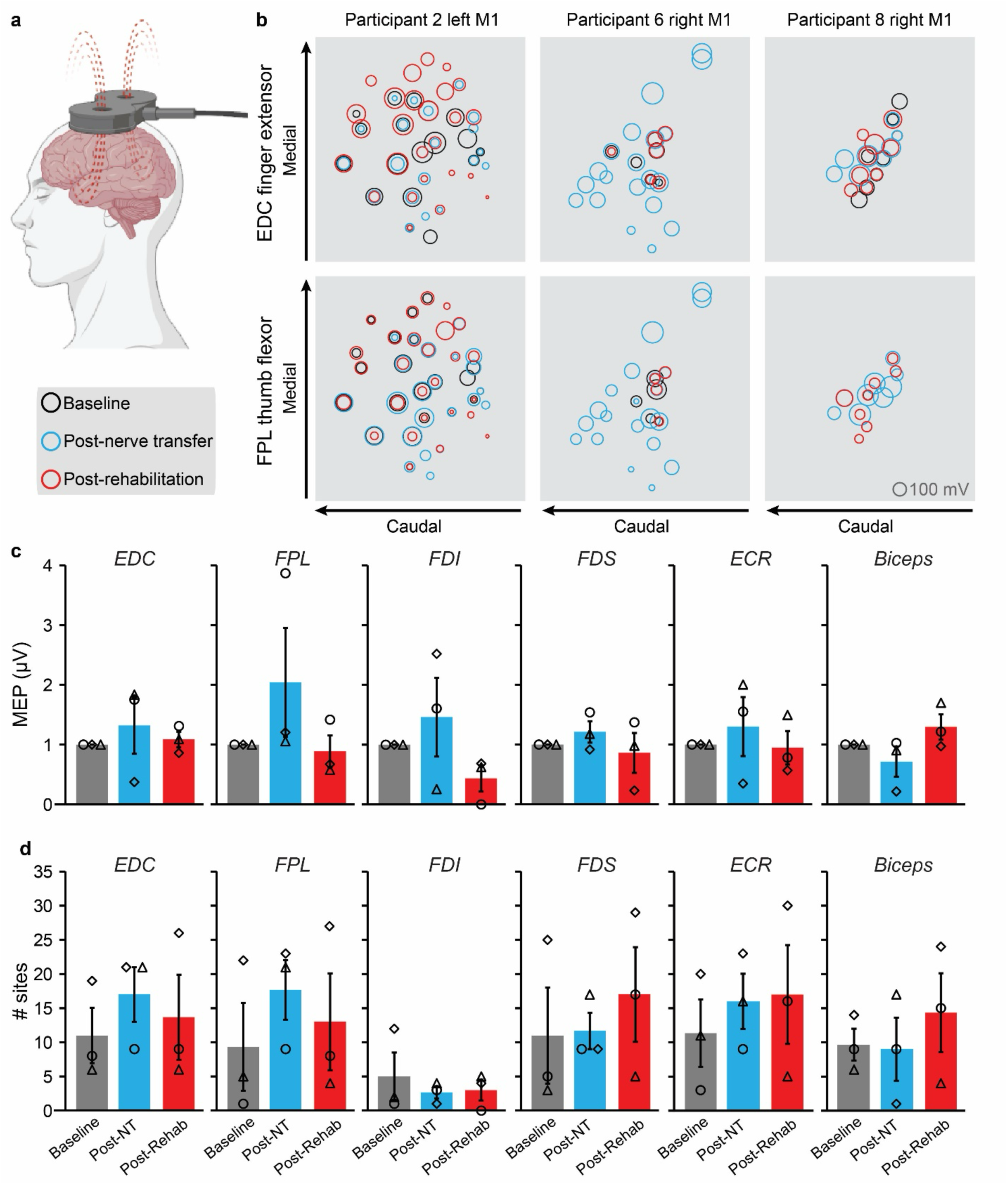
Transcranial magnetic stimulation (TMS) mapping of motor cortex shows dynamic changes in hand muscle representations. (**a**) TMS mapping schematic created with BioRender.com. (**b**) Maps of *EDC* and *FPL* responses illustrate dynamic motor map changes due to nerve transfer surgery and rehabilitation across individuals. (**c-d**) Nerve transfer surgery and robotic rehabilitation drove dynamic changes in hand muscle responsive sites with little effect on MEP amplitudes. (◊ = Participant 2 Right Arm, Δ= Participant 6, ○ = Participant 8). Data presented as mean ± SEM.

Pre-surgery mapping of the left arm was not possible. However, maps obtained 14 months post-surgery and again after robotics revealed patterns similar to those observed in the left motor cortex. After robotics, the left-arm motor map expanded, with a non-statistically significant increase in the number of responsive sites across all muscles (Table S8). MEP amplitudes moderately increased in *EDC, FPL, FDS, and ECR* and decreased in *FDI* and *Biceps* (Table S10).

At one year post-surgery, MEP amplitudes in Participant 6 remained unchanged relative to baseline for all muscles except *FDI* and *ECR*. Despite limited changes in excitability, the motor map moderately expanded after the nerve transfer surgery, with more responsive sites observed for all muscles (Table S7). The average MEP amplitude decreased in *FDI* at one year after surgery but mildly increased following six weeks of robotic training. Similarly, *Biceps* MEP amplitude also increased after the robotic intervention (Table S9).

In Participant 8, motor map size and MEP amplitude remained stable both one year after nerve transfer surgery and after six weeks of robotic rehabilitation. Nevertheless, six weeks of robotic training produced slight localized motor map expansion for *FDS*, *ECR*, and *Biceps*, characterized by an increased number of responsive sites and stronger MEP responses in *Biceps*.

## Discussion

We report preliminary evidence from three individuals with chronic tetraplegia demonstrating functional, kinematic, and cortical changes following robot-assisted rehabilitation after nerve transfer surgery. Across participants, the combined intervention appeared to support modest improvements in hand and arm function independent of age, chronicity, and baseline motor capacity as all individuals met the minimally important difference (MID) in at least one clinical strength measure, suggesting that pairing nerve transfer with robotic rehabilitation may drive meaningful recovery even many years post-injury.

In Participant 2, nerve transfer surgery supported increased hand strength and dexterity of the dominant right hand. Six weeks of robot-assisted rehabilitation further strengthened hand extensors and improved grasp range of motion, with modest increases in UEMS and GRASSP strength and prehension scores. Notably, performance with the non-dominant left hand declined on Box and Blocks after surgery, a pattern consistent with disruption of compensatory tenodesis grasp and reduction of spastic, passive tone that had previously facilitated hand opening and object stabilization as nerve transfer replaces passive spasticity with active voluntary control; however, if the newly recovered active output is initially weaker than prior passive tension, pinch strength may be reduced, leading to poorer performance on Box and Blocks and GRASSP prehension tasks. The pronounced rehabilitation-driven gains in right-hand extensors, despite a flexor-targeted nerve transfer, may also reflect reduced spastic resistance to opening, allowing greater aperture both actively and passively. TMS mapping paralleled these functional results: *EDC* representations mildly strengthened and expanded post-rehabilitation, indicating cortical reorganization that accommodated the novel peripheral circuitry.

Participants 6 and 8 exhibited more modest functional responses. Participant 6 experienced mild improvements primarily in kinematic measures, despite receiving transfers to the non-dominant limb. Participant 8 showed a decline in hand strength after surgery, but this was likely influenced by a tendon rupture unrelated to the study that occurred before robotic training. Both individuals nevertheless demonstrated small improvements following the robotic intervention, suggesting that paired surgical and robotic approaches may yield benefit even when confounding orthopedic events occur or when non-dominant limbs are treated.

Consistent with prior work, nerve transfer alone produced meaningful improvements in self-reported function and independence ^30,31^. Participant 2 demonstrated increases on the CUE questionnaire and in SCIM III self-care sub-scores, particularly in bathing, lower-body dressing, and grooming. Participants 6 and 8 also showed increases in CUE scores following surgery. These results highlight the substantial impact that restoration of even limited volitional control can have on quality of life. Robotic rehabilitation did not produce detectable changes on the SCIM III, likely due to its limited sensitivity to incremental distal arm improvements, though slight gains were captured by CUE ratings.

### Nerve transfer as a pathway for inclusion of AIS A individuals

A central implication of these findings is the potential for nerve transfer surgery to offer access to a therapeutic intervention for individuals living with AIS A injuries. Many emerging neuromodulation-based approaches, including transcutaneous spinal stimulation approaches ^5^, require some degree of spared ascending or descending conduction, demonstrated by residual sensation, subthreshold motor responses, or voluntary EMG activity. AIS A individuals lack these prerequisites and are therefore not eligible for such trials.

Nerve transfer surgery is not subject to this limitation as it is used to reinnervate distal muscles using donor fascicles from above-injury segments, thereby creating *new* volitional pathways that do not rely on spared spinal cord function. This makes nerve transfer viable for individuals with complete injuries who lack detectable descending input. In essence, nerve transfer creates a peripheral bypass for central nervous system plasticity that does not depend on preserved spinal pathways. Robotic rehabilitation then provides a high-dose, repeatable, task-specific training to strengthen these surgically established circuits and promote cortical reorganization.

In previous research, robotic rehabilitation alone in this population did not significantly impact any clinical measures, including UEMS ^14^. Together, nerve transfer and robotic rehabilitation may therefore broaden participation to a population of individuals who have traditionally been excluded from neuromodulation-based interventions. Our results suggest that even in AIS A injuries, meaningful functional improvements and cortical reorganization can occur through this combined approach, expanding opportunities for recovery research and ultimately for clinical rehabilitation.

Although moderate improvements were observed across behavioral, kinematic, and cortical plasticity, variability in individual responses highlights the need for studies with larger cohorts to determine the generalizability and magnitude of benefit that can be achieved with this combined approach. Additional participants will provide clarity as to which individuals stand to gain the most from nerve transfer surgery and how rehabilitation intensity, timing, and modality can be optimized to maximize outcomes.

Effective interventions for adults with chronic tetraplegia remain critically needed, especially for those with complete injuries (AIS A) who currently face limited options for improving function and independence. While this report includes only three individuals, it provides early evidence that pairing nerve transfer surgery with intensive robotic intervention may offer a promising avenue to enhance upper limb function and quality of life in adults with chronic cervical SCI, including those with the most severe injury classifications.

## Data Availability

Full dataset generated during this study will be available upon acceptance for publication in a publicly accessible repository at the Open Data Commons for Spinal Cord Injury (ODC-SCI, lab ID 110, Hollis lab).

## Acknowledgements

We thank Marissa Wuennemann, OTD, OTR/L; Lydia Currie; Devina Kumar, PhD; and Josh Silverstein for all their help and support conducting clinical evaluations and TMS sessions. We also thank Dylan Edwards, PhD, for his guidance in study development and Mahdi Safdarian, MD, for his editorial contributions to this study. This work was supported by the New York State Department of Health Spinal Cord Injury Research Program C34463GG and C37710GG, the Burke Foundation 103-001, the National Center for Advancing Translational Sciences (NCATS) grant UL1TR2384 of the Clinical and Translational Science Center at Weill Cornell Medical College, and the National Institutes of Health DP2 NS106663.

## SUPPLEMENT

**Table S1.** Clinical measures of hand function showed moderate improvement after nerve transfer surgery and mild improvement after 6 weeks of robotic training. Improvements from robotic training were largely driven by increases in hand strength rather than dexterity or sensation. Nerve transfer drove improved right hand performance on Box and Blocks for Participant 2. Robotic rehabilitation resulted in improved UEMS. GRASSP testing demonstrated effects of nerve transfer on motor function, with robotic rehabilitation supporting total prehension ability. Improvements in quality-of-life surveys were largely driven by nerve transfer, with some effect of robotic rehabilitation. UEMS: upper extremity motor score, GRASSP: Graded Redefined Assessment of Strength Sensibility and Prehension, 1: strength, 2: sensation, 3: prehension ability, 4: prehension performance, SCIM III: Spinal Cord Independence Measure self-care sub-score, CUE: Capabilities of Upper Extremity Instrument single arm score

| *Outcome Measures* | *Participant 2 Right Hand* |  |  | *Participant 6* |  |  | *Participant 8* |  |  |
| --- | --- | --- | --- | --- | --- | --- | --- | --- | --- |
|  | Baseline | Post-NT | Post-Rehab | Baseline | Post-NT | Post-Rehab | Baseline | Post-NT | Post-Rehab |
| *Box and Blocks* | 17 | 25 | 23 | 1 | 1 | 2 | 5 | 5 | 8 |
| *UEMS* | 14 | 15 | 18 | 8 | 11 | 11 | 14 | 11 | 16 |
| *GRASSP Strength* | 13 | 23 | 25 | 12 | 25 | 22 | 19 | 17 | 21 |
| *GRASSP Sensation* | 7 | 7 | 7 | 6 | 4 | 3 | 8 | 10 | 10 |
| *GRASSP Grip* | 2 | 6 | 5 | 2 | 0 | 0 | 4 | 7 | 7 |
| *GRASSP Prehension* | 8 | 7 | 10 | 3 | 2 | 1 | 4 | 6 | 8 |
| *SCIM III self-care* | 5 | 9 | 9 | 4 | 3 | 3 | 8 | 5 | 5 |
| *CUE* | 34 | 42 | 38 | 24 | 19 | 25 | 29 | 27 | 36 |
| *MAS Wrist* | 1 | 0 | 0 | 2 | 0 | 1 | 0 | 0 | 0 |

**Table S2.** Clinical measures of hand function showed variable effects after nerve transfer surgery and mild improvement after 6 weeks of robotic training. Some decline in performance was seen due to the lack of compensatory strategies that were able to be employed after the nerve transfer surgery. Improvements from robotic training were largely driven by increases in hand strength rather than dexterity or sensation. UEMS: upper extremity motor score, GRASSP: Graded Redefined Assessment of Strength Sensibility and Prehension, 1: strength, 2: sensation, 3: prehension ability, 4: prehension performance, SCIM III: Spinal Cord Independence Measure self-care sub-score, CUE: Capabilities of Upper Extremity Instrument single arm score

| Outcome Measures | Participant 2 Left Hand |  |  | MID |  |  |
| --- | --- | --- | --- | --- | --- | --- |
|  | Baseline | Post-NT | Post-Rehab | Baseline → Post-NT | Post-NT → Post-Rehab | Baseline → Post-Rehab |
| Box and Blocks | 15 | 6 | 4 | -9 | -2 | 11 |
| UEMS MID= 2 | 13 | 14 | 13 | 1 | -1 | 0 |
| GRASSP Strength MID= 5 | 13 | 22 | 18 | 9 | -4 | 5 |
| GRASSP Sensation MID= 4 | 7 | 7 | 4 | 0 | -3 | -3 |
| GRASSP Grip MID= 4 | 2 | 1 | 3 | -1 | 2 | 1 |
| GRASSP Prehension MID= 3 | 6 | 2 | 3 | -4 | 1 | -3 |
| SCIM III self-care MID= 1.15 | 5 | 7 | 8 | 2 | 1 | 3 |
| CUE | 28 | 31 | 34 | 3 | 3 | 6 |
| MAS Wrist | 0 | 0 | 0 | 0 | 0 | 0 |

**Table S3.** Kinematic measures of hand function from InMotion HAND/ARM robot showed mild improvement after nerve transfer surgery and six weeks of robotic training. Grasp and release force and grasp active range of motion improved on the Bionik Robot for Participant 2. Healthy control values provided by Bionik. ^†^Smaller values indicate better performance.

| Outcome | Healthy | Participant 2 Right Hand |  |  | Participant 6 |  |  | Participant 8 |  |  |
| --- | --- | --- | --- | --- | --- | --- | --- | --- | --- | --- |
| Measures | Control | Baseline | Post-NT | Post-Rehab | Baseline | Post-NT | Post-Rehab | Baseline | Post-NT | Post-Rehab |
| Hand Release AROM | 7.8 cm | 4.07 cm | 4.09 cm | 4.08 cm | 4.18 cm | 4.18 cm | 4.26 cm | 4.06 cm | 4.08 cm | 4.08 cm |
| Hand Grasp AROM<sup>†</sup> | 4.2 cm | 4.63 cm | 4.28 cm | 4.23 cm | 6.82 cm | 4.42 cm | 4.39 cm | 3.95 cm | 3.96 cm | 4.32 cm |
| Hand Release Stabilization | 6.7 cm | 4.36 cm | 4.35 cm | 4.5 cm | 4.39 cm | 4.39 cm | 4.83 cm | 4.37 cm | 4.38 cm | 4.38 cm |
| Hand Release Force | 14 N | 0.5 N | 0.3 N | 2 N | 1 N | 1.9 N | 1.9 N | 0.3 N | 0.1 N | 0.1 N |
| Hand Grasp Stabilization<sup>†</sup> | 4.5 cm | 7.22 cm | 6.71 cm | 6.68 cm | 7.39 cm | 7.15 cm | 6.86 cm | 5.75 cm | 6.2 cm | 6.85 cm |
| Hand Grasp Force | 34.7 N | 6.6 N | 12.3 N | 12.4 N | 4.8 N | 7.9 N | 10.6 N | 22.6 N | 17.1 N | 10.3 N |

**Table S4.** Kinematic measures of hand function from InMotion HAND/ARM robot showed mild improvement after six weeks of robotic training. Grasp and release force and release active range of motion improved on the Bionik Robot. Healthy control values provided by Bionik. ^†^Smaller values indicate better performance.

| *Outcome Measures* | **Healthy** | *Participant 2 Left Hand* |  |
| --- | --- | --- | --- |
|  | **Control** | Post-NT | Post-Rehab |
| *Hand Release AROM* | **7.8 cm** | 4.12 cm | 4.17 cm |
| *Hand Grasp AROM<sup>†</sup>* | **4.2 cm** | 4.37 cm | 4.40 cm |
| *Hand Release Stabilization* | **6.7 cm** | 4.39 cm | 4.42 cm |
| *Hand Release Force* | **14 N** | 0.8 N | 1 N |
| *Hand Grasp Stabilization<sup>†</sup>* | **4.5 cm** | 7.39 cm | 7.03 cm |
| *Hand Grasp Force* | **34.7 N** | 5.5 N | 9.0 N |

**Table S5.** Kinematic measures of arm function from InMotion HAND/ARM robot. Arm function kinematic measures remained largely unchanged from baseline, indicating the nerve transfer did not result in any loss of function of the upper arm. Minimal change was expected since both participants had good arm function before receiving the nerve transfer surgery. Healthy control values provided by Bionik. ^†^Smaller values indicate better performance.

| Outcome Measures | Healthy Control | Participant 2 Right Hand |  |  | Participant 6 |  |  | Participant 8 |  |  |
| --- | --- | --- | --- | --- | --- | --- | --- | --- | --- | --- |
|  |  | Baseline | Post-NT | Post-Rehab | Baseline | Post-NT | Post-Rehab | Baseline | Post-NT | Post-Rehab |
| Circle Size | **100.53 cm** | 85.61 cm | 87.9 cm | 88.55 cm | 58.47 cm | 80.73 cm | 76.6 cm | 77.16 cm | 82.95 cm | 79.18 cm |
| Circle Symmetry | **0.834** | 0.925 | 0.873 | 0.906 | 0.629 | 0.641 | 0.672 | 0.581 | 0.86 | 0.84 |
| Target Accuracy<sup>†</sup> | **0.85 cm** | 0.92 cm | 0.88 cm | 0.91 cm | 4.33 cm | 1.96 cm | 0.8 cm | 0.84 cm | 0.84 cm | 0.88 cm |
| Movement Efficiency | **75%** | 88% | 90% | 88% | 40% | 52% | 67% | 74% | 85% | 84% |
| Isometric Hold Distance<sup>†</sup> | **2.4 cm** | 4.81 cm | 4.48 cm | 4.11 cm | 9.48 cm | 8.5 cm | 8.32 cm | 6.61 cm | 5.4 cm | 5.35 cm |
| Isometric Hold Force | **26 N** | 20.1 N | 20.7 N | 21.3 N | 17.6 N | 15.2 N | 13.7 N | 20.3 N | 20.2 N | 21 N |
| Displacement Against Resistance | **13 cm** | 12.8 cm | 13.06 cm | 13.09 cm | 6.56 cm | 11.52 cm | 11.31 cm | 11.87 cm | 13.06 cm | 10.34 cm |
| Resistance Force | **26 N** | 26 N | 26.7 N | 26.8 N | 18 N | 22.9 N | 25.4 N | 27.2 N | 28.2 N | 22.2 N |

**Table S6.**
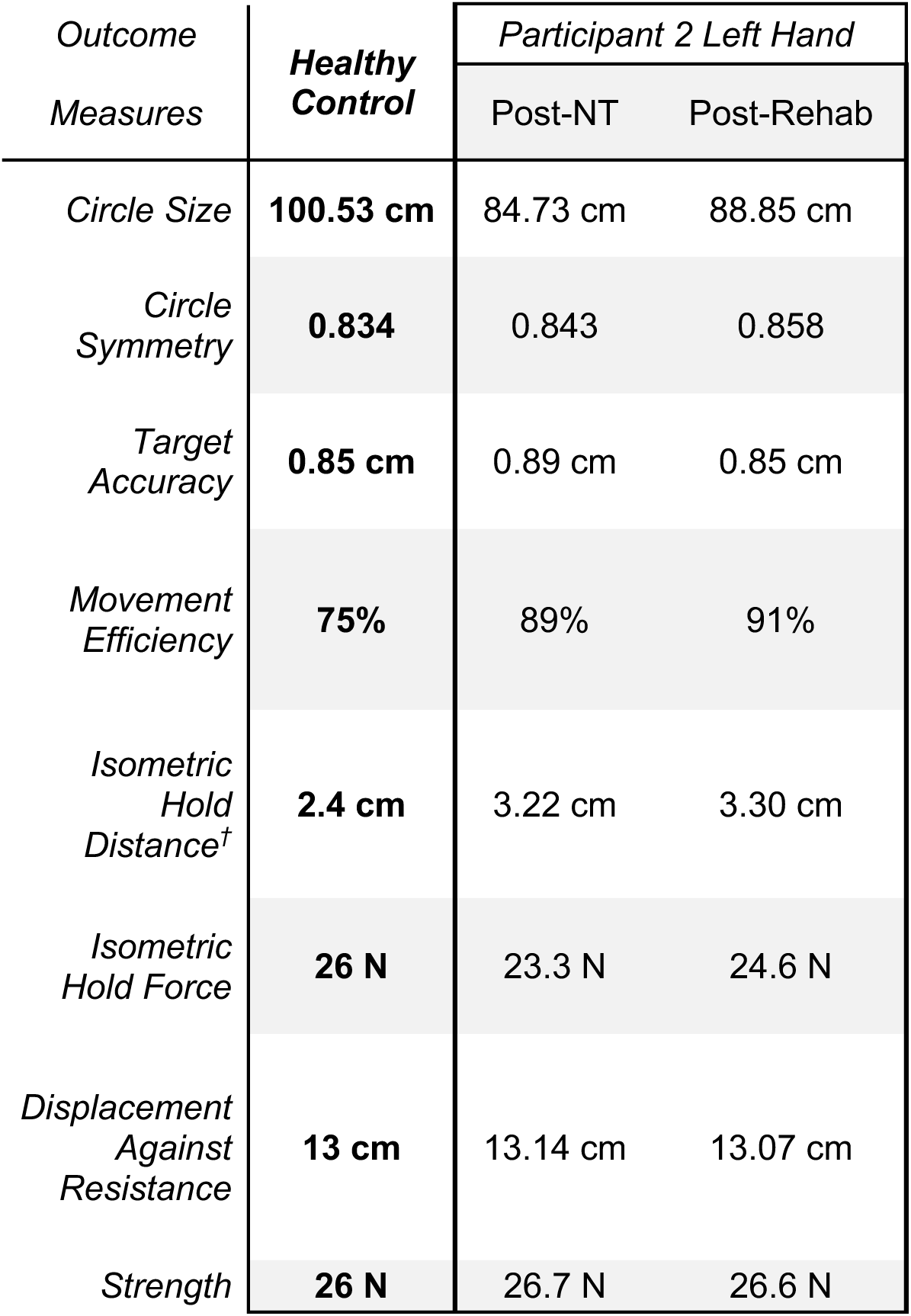
Kinematic measures of arm function from InMotion HAND/ARM robot. Arm function kinematic measures remained largely unchanged from the start of rehab, indicating the hand therapy had minimal effect on the arm function. Healthy control values provided by Bionik. ^†^Smaller values indicate better performance.

| *Outcome Measures* | ***Healthy Control*** | *Participant 2 Left Hand* |  |
| --- | --- | --- | --- |
|  |  | Post-NT | Post-Rehab |
| *Circle Size* | **100.53 cm** | 84.73 cm | 88.85 cm |
| *Circle Symmetry* | **0.834** | 0.843 | 0.858 |
| *Target Accuracy* | **0.85 cm** | 0.89 cm | 0.85 cm |
| *Movement Efficiency* | **75%** | 89% | 91% |
| *Isometric Hold Distance<sup>†</sup>* | **2.4 cm** | 3.22 cm | 3.30 cm |
| *Isometric Hold Force* | **26 N** | 23.3 N | 24.6 N |
| *Displacement Against Resistance* | **13 cm** | 13.14 cm | 13.07 cm |
| *Strength* | **26 N** | 26.7 N | 26.6 N |

**Table S7.**
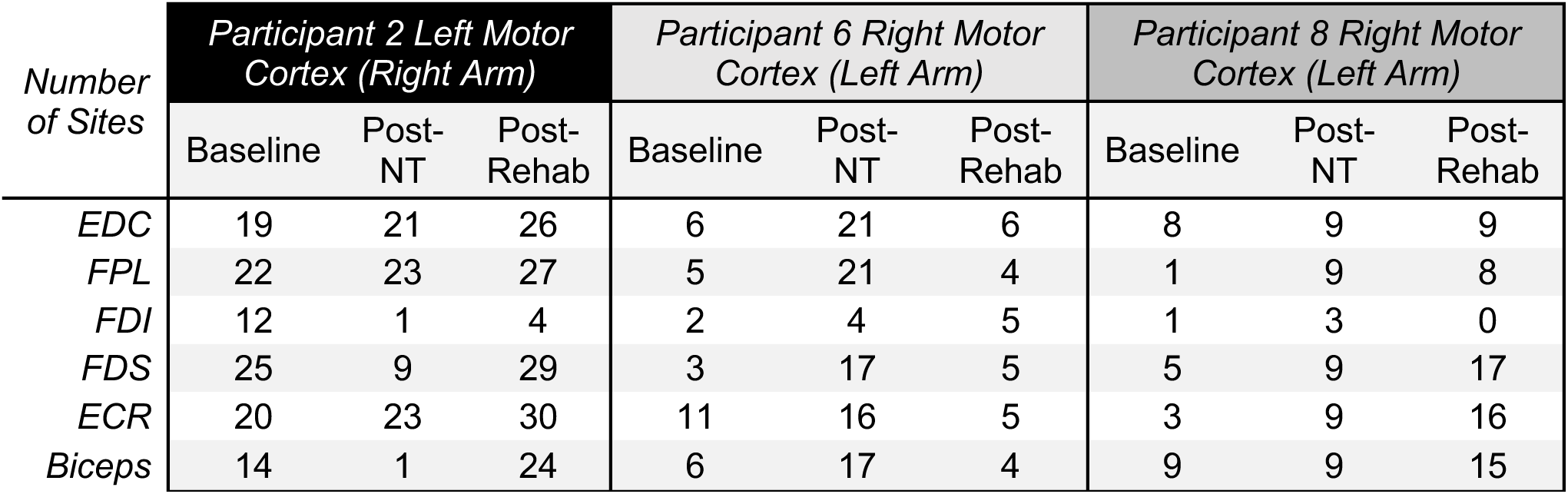
Number of sites of the left and right motor cortex for 6 upper extremity muscles. Motor map size changed to accommodate new circuits one year after nerve transfer surgery and after six weeks of robotic training.

| Number of Sites | Participant 2 Left Motor Cortex (Right Arm) |  |  | Participant 6 Right Motor Cortex (Left Arm) |  |  | Participant 8 Right Motor Cortex (Left Arm) |  |  |
| --- | --- | --- | --- | --- | --- | --- | --- | --- | --- |
|  | Baseline | Post-NT | Post-Rehab | Baseline | Post-NT | Post-Rehab | Baseline | Post-NT | Post-Rehab |
| *EDC* | 19 | 21 | 26 | 6 | 21 | 6 | 8 | 9 | 9 |
| *FPL* | 22 | 23 | 27 | 5 | 21 | 4 | 1 | 9 | 8 |
| *FDI* | 12 | 1 | 4 | 2 | 4 | 5 | 1 | 3 | 0 |
| *FDS* | 25 | 9 | 29 | 3 | 17 | 5 | 5 | 9 | 17 |
| *ECR* | 20 | 23 | 30 | 11 | 16 | 5 | 3 | 9 | 16 |
| *Biceps* | 14 | 1 | 24 | 6 | 17 | 4 | 9 | 9 | 15 |

**Table S8.**
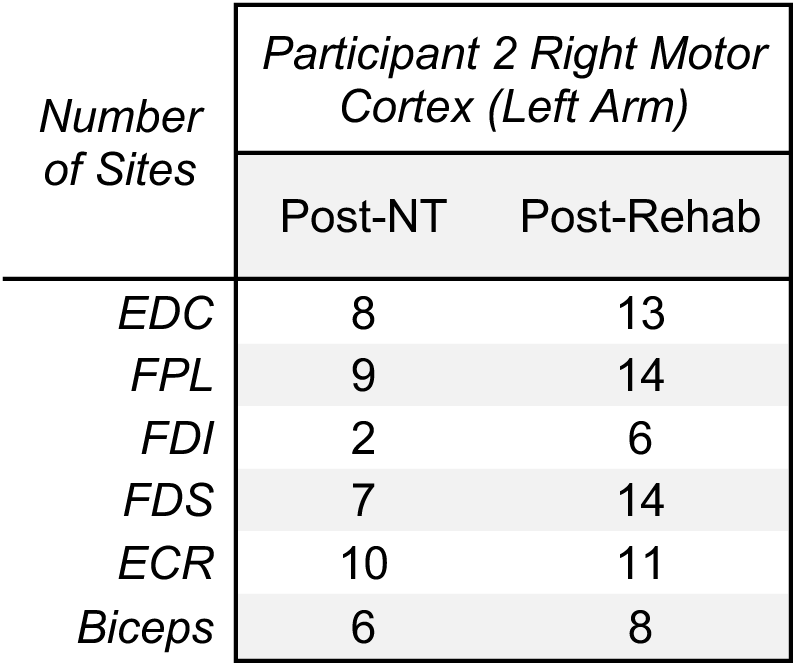
Number of sites of the right motor cortex for 6 upper extremity muscles. Motor map size changed to accommodate new circuits after six weeks of robotic training.

| Number of Sites | Participant 2 Right Motor Cortex (Left Arm) |  |
| --- | --- | --- |
|  | Post-NT | Post-Rehab |
| *EDC* | 8 | 13 |
| *FPL* | 9 | 14 |
| *FDI* | 2 | 6 |
| *FDS* | 7 | 14 |
| *ECR* | 10 | 11 |
| *Biceps* | 6 | 8 |

**Table S9.**
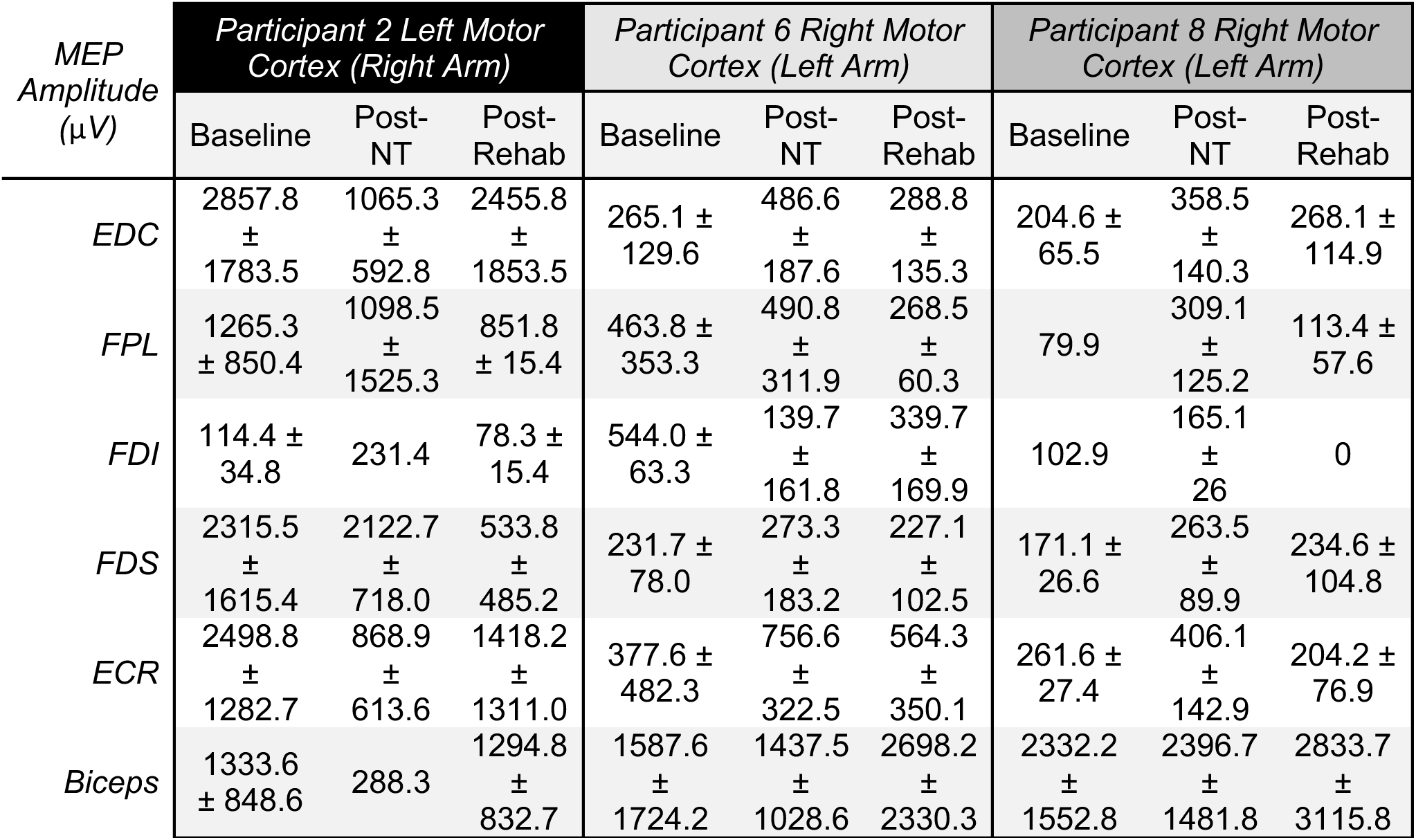
MEP Amplitude (µV ± SD) of the left and right motor cortex for 6 upper extremity muscles. Motor strength changed to accommodate new circuits one year after nerve transfer surgery and after six weeks of robotic training.

**Table S10.**
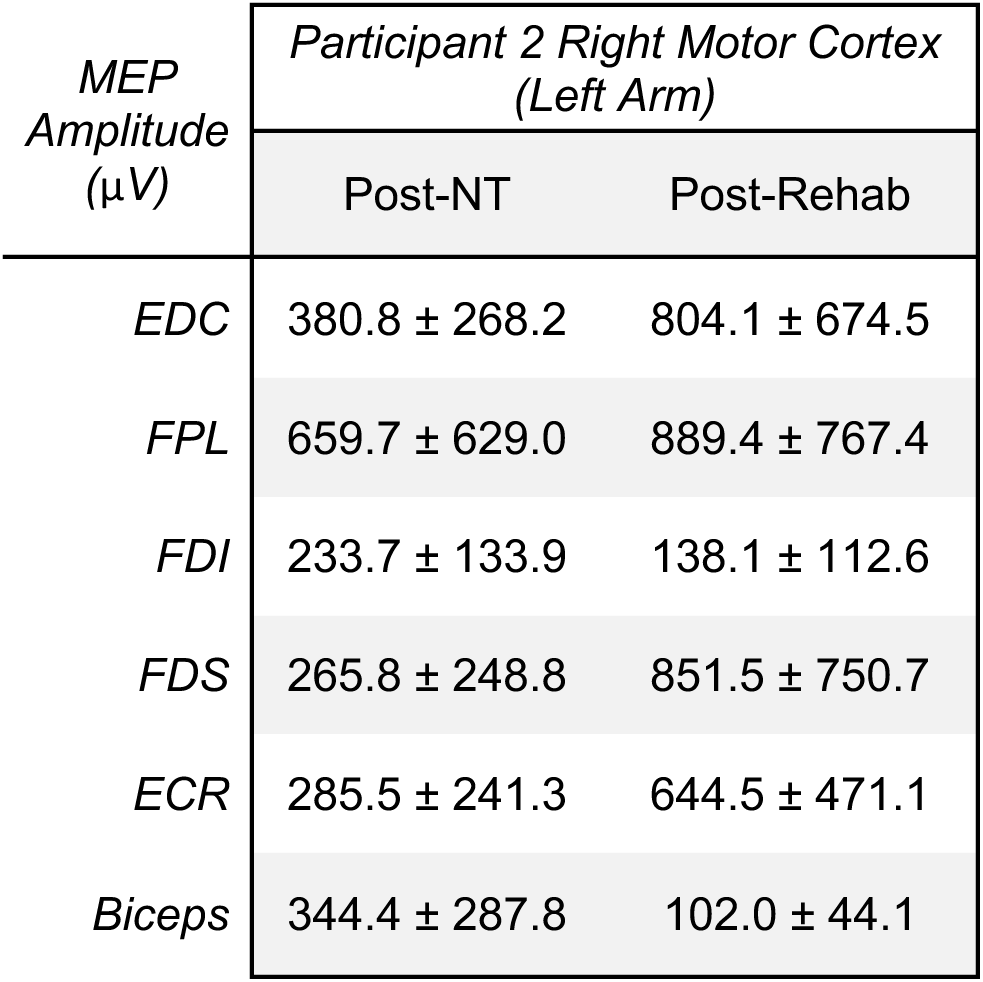
MEP Amplitude (µV ± SD) of the right motor cortex for 6 upper extremity muscles. Motor strength changed to accommodate new circuits after six weeks of robotic training.

**Figure S1.**
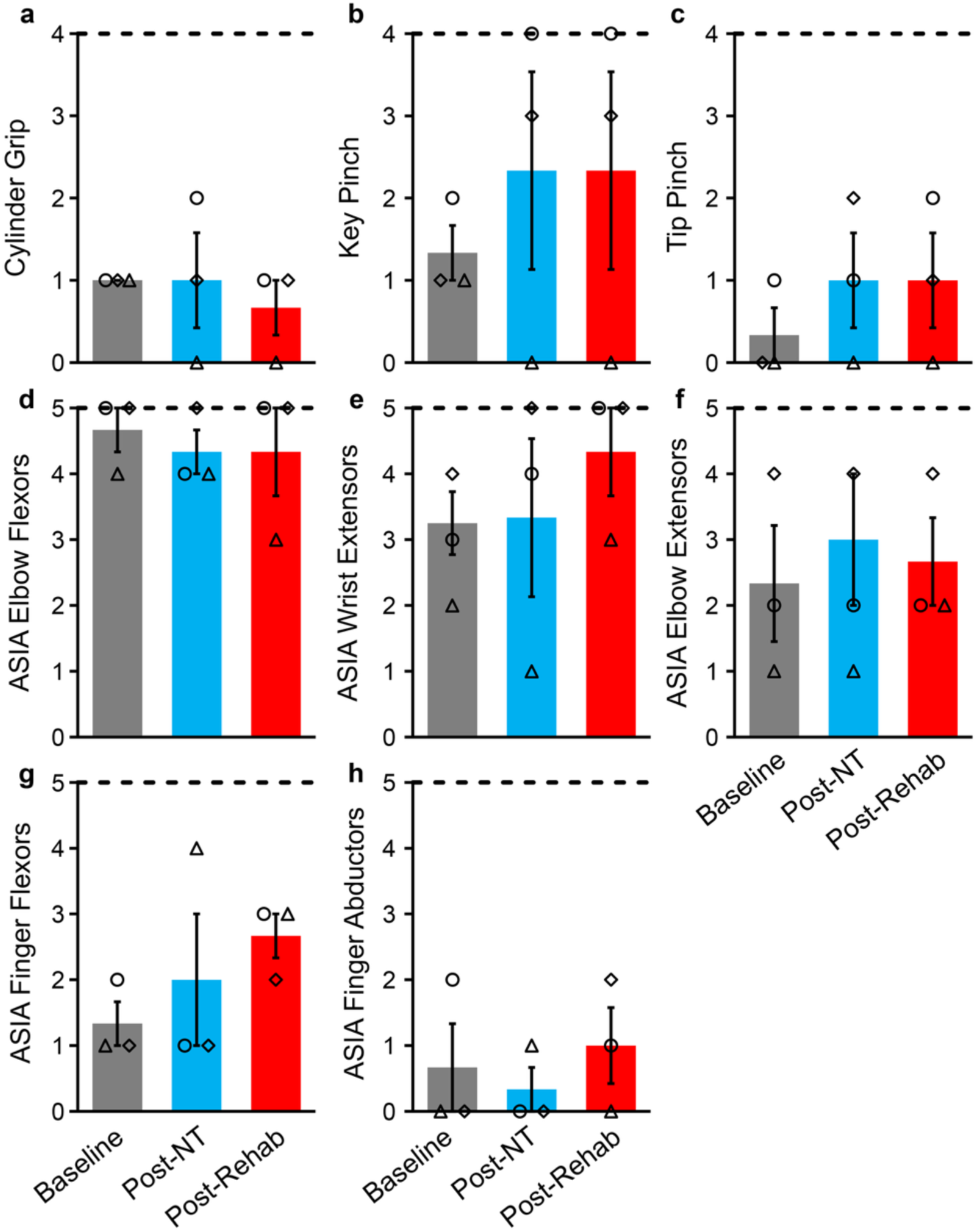
Additional clinical outcome scores. (**a-c**) Nerve transfer surgery drove improvements in grips focused on the index finger and thumb on GRASSP Grip test. (**d-h**) Improvements in individual UEMS were driven by increased strength in the wrist extensors, finger flexors, and finger abductors. (**◊ =** Participant 2 Right Arm, Δ= Participant 6, **○** = Participant 8).

**Figure S2.**
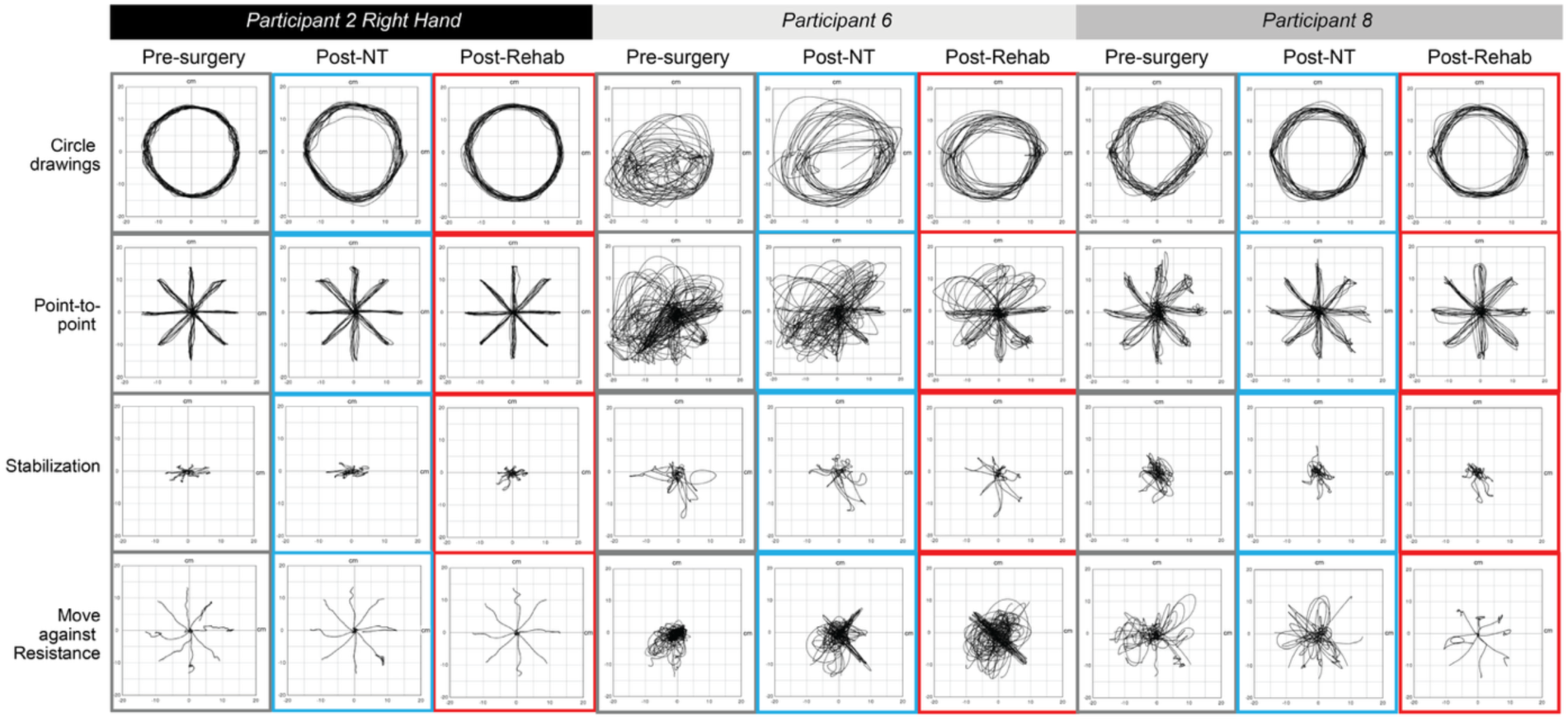
Kinematic traces of InMotion HAND/ARM evaluations. No loss of function in the arm was seen after nerve transfer surgery, and movement quality improved after six weeks of rehabilitation.

**Figure S3.**
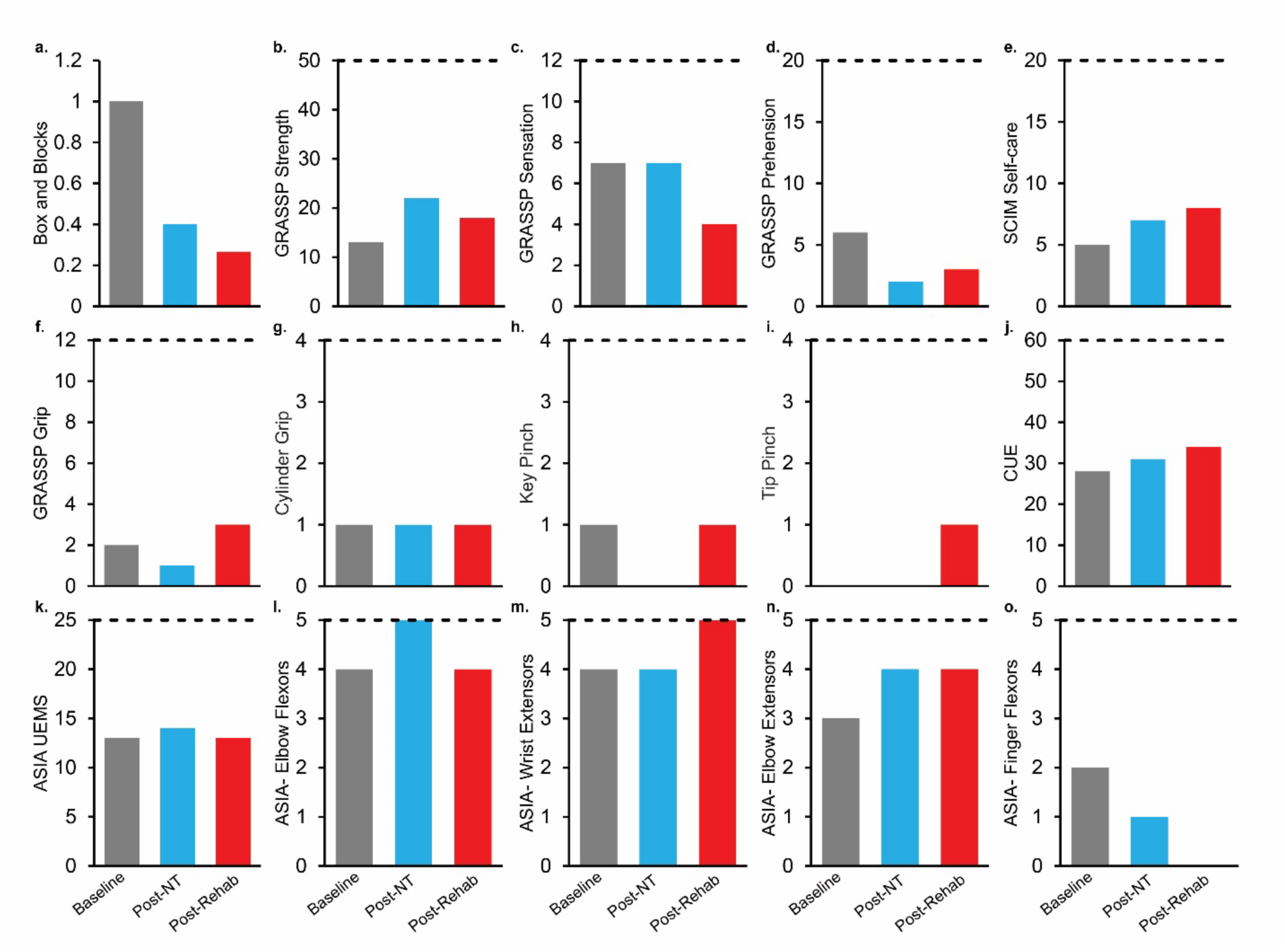
Clinical outcome scores show minimal improvement in hand function after nerve transfer and robotic rehabilitation in Participant 2’s left arm. Some decline in functional performance in box and blocks was seen due to the inability to use previously learned compensatory motions.

**Figure S4.**
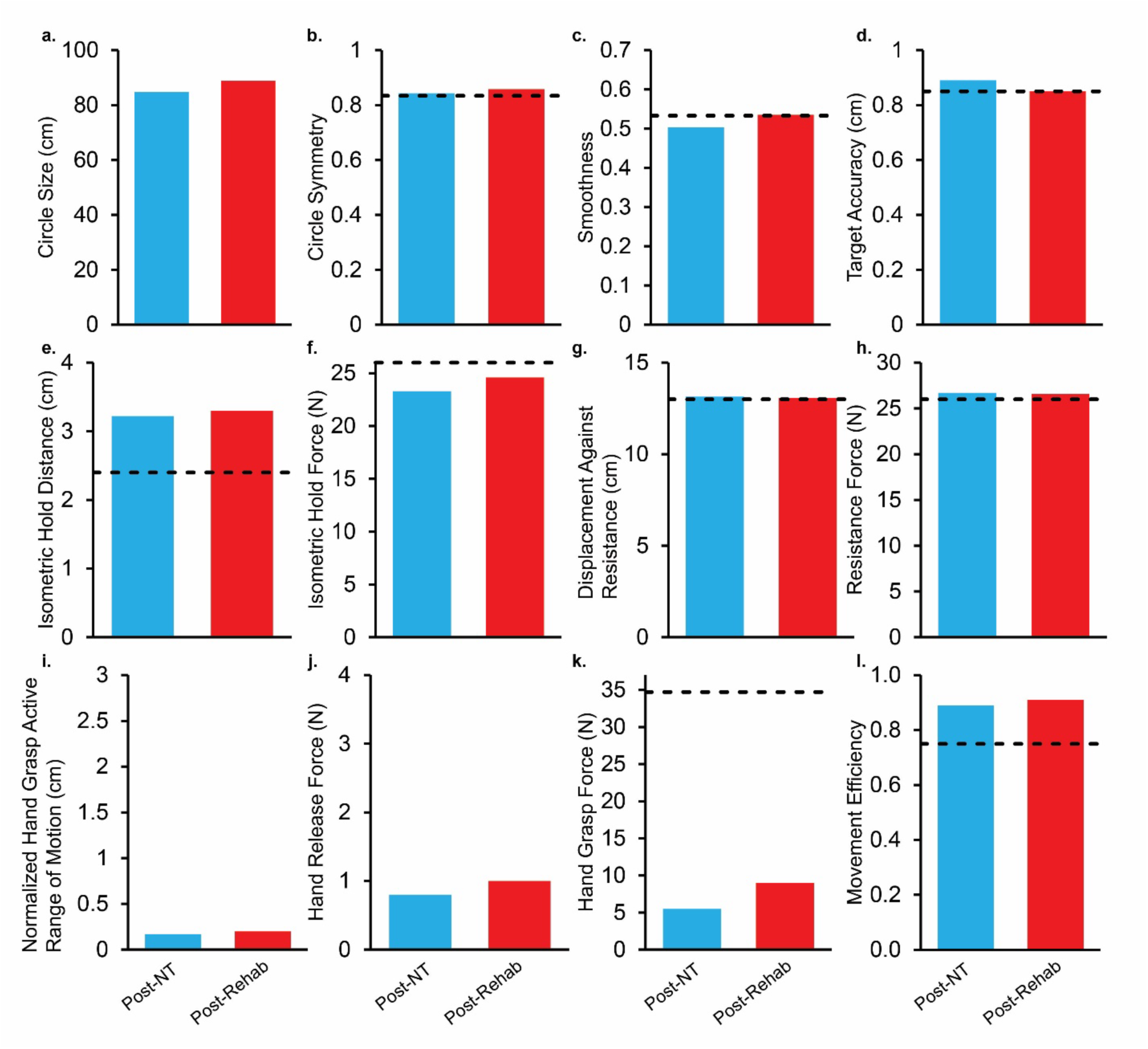
Measures from InMotion HAND/ARM robot show no loss of function from the nerve transfer surgery and improved strength of hand movements in Participant 2’s left arm. Target values represented by heavy dashed lines.(**a-h, l**) Minimal changes in kinematics after nerve transfer surgery and robotic training. (**j**, **k**) Robotic rehabilitation supported the restoration of hand grasp and release force

**Figure S5.**
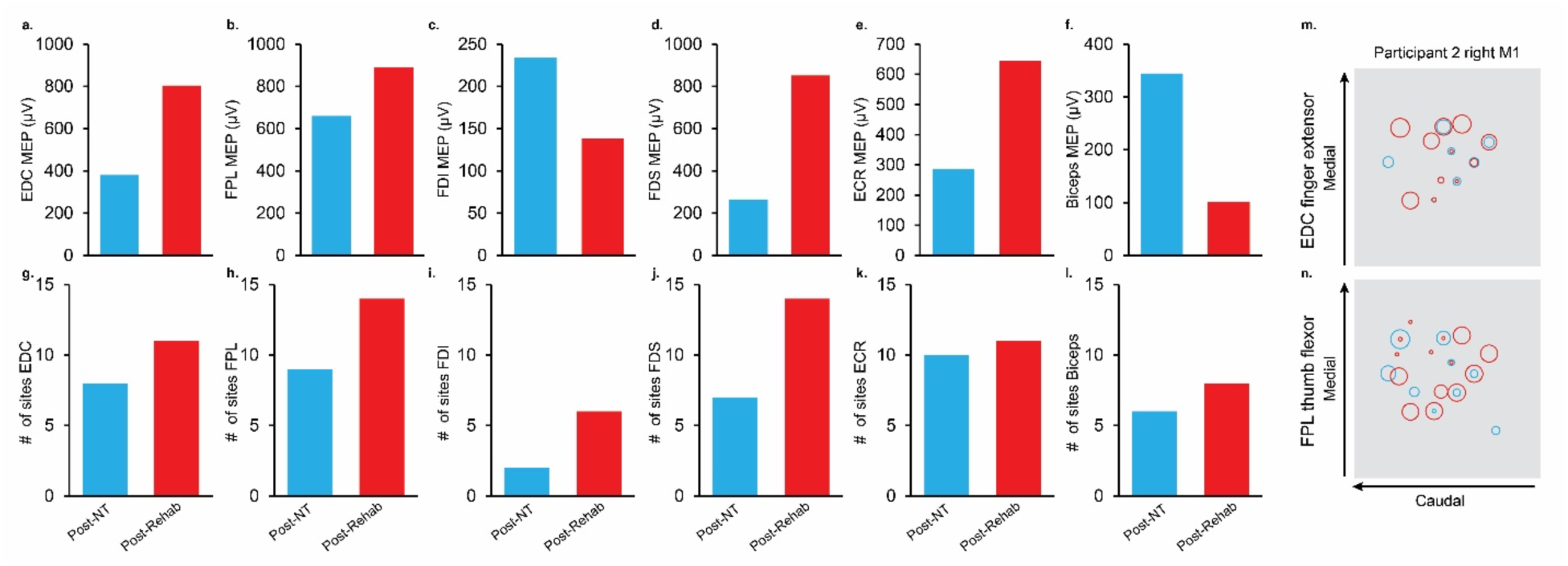
Transcranial magnetic stimulation mapping of motor cortex shows rehabilitation-mediated expansion of hand muscle representations in Participant 2’s left arm. (**a, b, d, e**) Rehabilitation drove increase in MEP amplitude in the EDC, FPL, FDS, and ECR. (**g-l**) Rehabilitation drove increases in total number of sites for all muscles. (**m-n**) Maps of *EDC* and *FPL* responses illustrate dynamic motor map changes due to nerve transfer surgery and rehabilitation.

